# The relationship between maternal psychopathology and offspring incontinence and constipation at school age: a prospective cohort study

**DOI:** 10.1101/2022.12.07.22283220

**Authors:** Gemma Sawyer, Jon Heron, Carol Joinson

**Author notes:** Correspondence to Gemma Sawyer at Oakfield House, Oakfield Grove, Clifton, Bristol, BS8 2BN, England.

## Abstract

**Background:** Maternal depression and anxiety may increase the risk of offspring incontinence; however, current evidence is unable to draw causal inferences. This study aimed to examine prospective associations between maternal psychopathology and offspring incontinence/constipation and examine evidence for causal intra-uterine effects.

**Methods:** The study used data from 6,489 children from the Avon Longitudinal Study of Parents and Children. Mothers provided data on depression and anxiety (antenatal and postnatal) and their child’s incontinence (daytime wetting, bedwetting, soiling) and constipation at age 7. We used multivariable logistic regression to examine evidence for independent effects of maternal depression/anxiety on offspring incontinence/constipation and explore a critical/sensitive period of exposure. A negative control design was utilised to examine evidence causal intra-uterine effects.

**Results:** Postnatal maternal psychopathology was associated with an increased risk of offspring incontinence and constipation (e.g. postnatal anxiety and daytime wetting OR: 1.53; 95% CI: 1.21-1.94), and data were consistent with a critical period model. There was evidence for an independent effect of maternal anxiety. Antenatal maternal psychopathology was associated with constipation (e.g. antenatal anxiety OR: 1.57; 95% CI: 1.25-1.98), but there was no evidence for a causal intra-uterine effect.

**Limitations:** Attrition and maternal reports without use of established diagnostic criteria for incontinence/constipation are potential limitations.

**Conclusions:** Children exposed to maternal postnatal psychopathology had a greater risk of incontinence/constipation, and maternal anxiety had stronger associations than depression.

## Introduction

Maternal psychopathology has been identified as a key risk factor for multiple adverse child outcomes, including cognitive and emotional development (Beck, 1998), social engagement, stress reactivity (Feldman et al., 2009), internalising and externalising behaviour (Goodman et al., 2011), poorer infant growth (Rahman et al., 2004), and physical health during childhood (National Research Council, 2009; Raposa et al., 2014). There is also some evidence that exposure to maternal psychopathology is associated with an increased risk of childhood incontinence and constipation.

Incontinence is common in childhood and most cases arise from functional impairments in the bladder and/or bowel rather than organic causes. Daytime urinary incontinence and bedwetting (enuresis) are estimated to affect around 8% and 15% of 7-year-olds respectively (Butler et al., 2005; Joinson et al., 2006b). Soiling (encopresis) is also common, with around 6.8% of children affected at age 7 (including 1.4% who soil once a week or more) (Joinson et al., 2006a). Over 80% of cases of childhood soiling are a consequence of chronic constipation, whilst around 20% have no underlying constipation (non-retentive) (Bongers et al., 2006). Despite the common belief that incontinence resolves with age, it can persist into adolescence, with prevalence estimates of around 3% for daytime wetting and 2.5% for bedwetting (Heron et al., 2017). Moreover, among children with soiling, 15% are estimated to continue to be affected at age 18, whilst 20% with constipation continue to experience symptoms at age 16 (Bongers et al., 2006). Incontinence has adverse consequences for children and adolescents, including low self-esteem, difficult peer relationships, and depressive symptoms (Grzeda et al., 2017; Thibodeau et al., 2013). It is, therefore, important to identify modifiable risk factors, possibly including maternal psychopathology, that could increase the risk of children having problems attaining and maintaining continence.

Studies have found evidence that exposure to maternal psychopathology could be a contributor to childhood incontinence and constipation. Cross-sectional studies have reported higher levels of psychopathology in mothers of children with bedwetting (Durmaz et al., 2017), constipation (Appak et al., 2017), and soiling (Akdemir et al., 2015) compared to mothers of children without these problems. Clinical studies have found higher anxiety scores in mothers of children with enuresis compared to controls without incontinence (Naitoh et al., 2012). These studies are unable to disentangle the direction of association between maternal psychopathology and childhood incontinence because reverse causality is plausible, e.g. mothers of children with bedwetting had decreased state anxiety following their child’s successful treatment (Naitoh et al., 2012). In addition, these studies failed to adequately adjust for confounders.

A prospective study found that lifetime parental psychopathology is associated with daytime wetting and bedwetting in offspring but did not adjust for confounders (Kessel et al., 2017). Other prospective studies have found that maternal psychopathology in the antenatal and postnatal periods is associated with children’s incontinence up to age 9 (Joinson et al., 2019, 2009, 2008). These studies leave several questions unanswered. Firstly, they did not examine whether maternal depression and anxiety have independent effects on offspring incontinence/constipation. Although maternal anxiety and depression are highly comorbid, they may have different effects on offspring development (Glover, 2014). Secondly, it is unclear whether exposure to maternal psychopathology in the antenatal or postnatal period is more important in determining the risk of offspring incontinence/constipation. It has been suggested that intra-uterine exposure to maternal psychopathology is associated with alterations in the foetal hypothalamic-pituitary-adrenal (HPA) axis, which can affect child development(Glover, 2014), whilst postnatal exposure could affect child development through adverse effects on parenting behaviour. Thirdly, it is unclear whether intra-uterine exposure to maternal psychopathology has a causal effect on offspring incontinence/constipation. Mother’s partner’s exposures assessed during pregnancy can be used as negative controls in observational studies to improve causal inference regarding effects of antenatal exposures on offspring because they have similar confounding structures to maternal antenatal exposures, but no plausible biological link with offspring outcomes. If maternal, but not partner, exposures are associated with offspring outcomes, this would provide evidence of a possible causal intra-uterine effect. Alternatively, if both mother and partner exposures are associated with offspring outcomes, confounding by shared environmental or genetic factors is the more likely explanation (Lipsitch et al., 2010).

The current study uses data from a large UK birth cohort to examine the relationship between maternal psychopathology (depression and anxiety) and offspring incontinence (daytime wetting, bedwetting, soiling) and constipation at 7 years. The objectives are to examine (i) evidence for independent effects of maternal depression and anxiety on these offspring outcomes, (ii) whether there is a critical or sensitive period of exposure to maternal psychopathology in the antenatal or postnatal period, and (iii) evidence for a causal intra-uterine effect of maternal psychopathology on offspring incontinence and constipation with a negative control design.

## Method

### Study population

The Avon Longitudinal Study of Parents and Children (ALSPAC) is a UK population-based birth cohort which recruited pregnant women residing in the former Avon region with expected delivery dates between 1^st^ April 1991 and 31^st^ December 1992. The initial cohort included 14,541 pregnancies, resulting in 13,988 children who were alive at 1 year of age. Full details of the ALSPAC methods have been previously outlined (Boyd et al., 2013; Fraser et al., 2013). The study website contains details of available data through a fully searchable data dictionary and variable search tool (http://www.bristol.ac.uk/alspac/researchers/our-data/). Ethical approval for the study was obtained from the ALSPAC Ethics and Law Committee and the Local Research Ethics Committees.

Informed consent for the use of data collected via questionnaires and clinics was obtained from participants following the recommendations of the ALSPAC Ethics and Law Committee at the time. The current study uses a complete case sample of 6,489 participants who provided data on all relevant variables (Figure 1).

**Figure 1.**
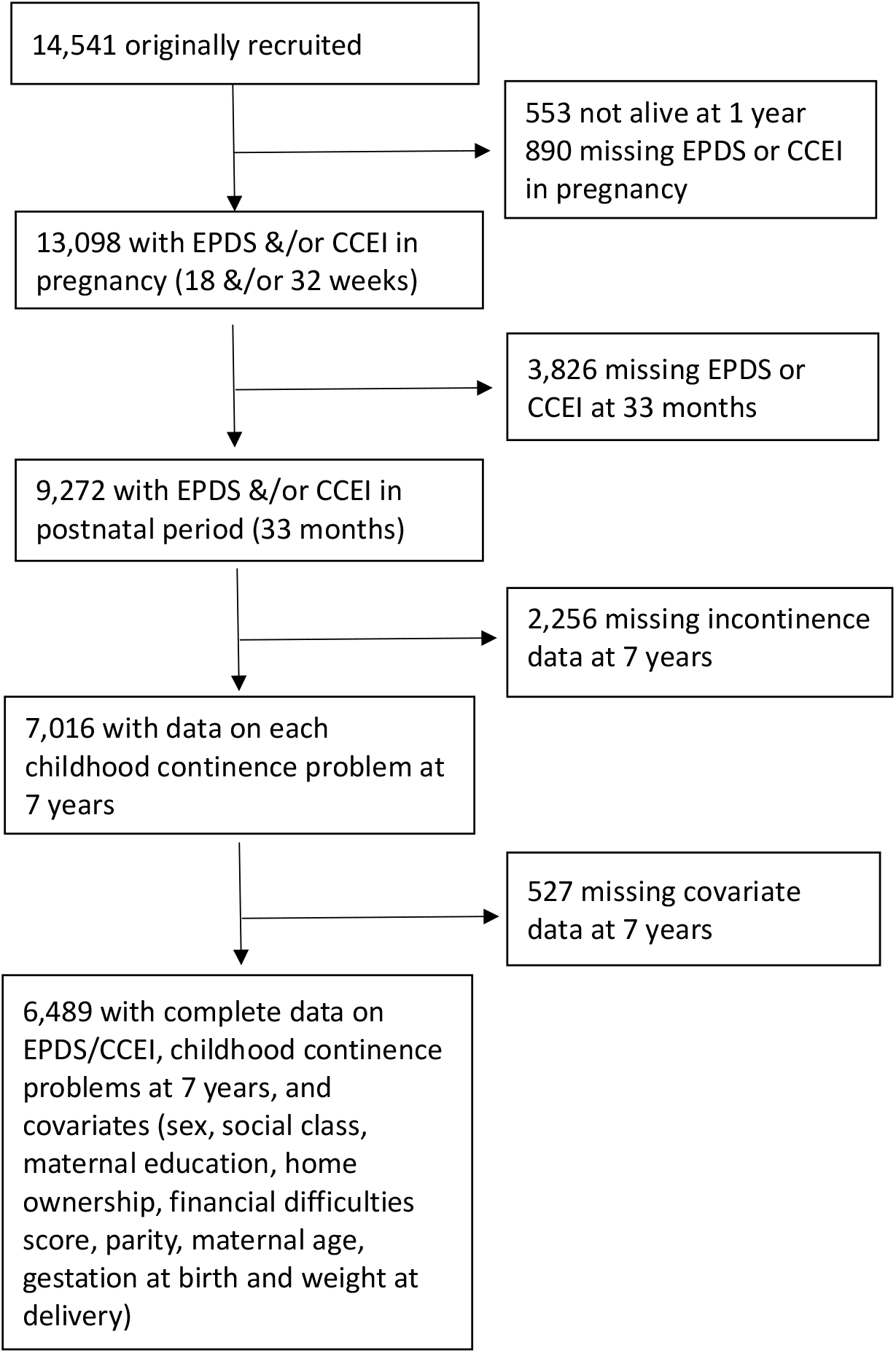
Flow chart showing the participants included in the study

### Exposures: maternal antenatal and postnatal depression and anxiety

At 18- and 32-weeks’ gestation and 33 months postnatal, mothers were invited to complete the Edinburgh Postnatal Depression Scale (EPDS), a validated measure for screening depression during and following pregnancy (Cox et al., 1987). Scores of 13 or higher are often used to indicate probable depression (Evans et al., 2001). We categorised mothers as ‘depressed’ or ‘not depressed’ in the antenatal and postnatal periods using this threshold. Mothers also completed the anxiety subscale of the Crown Crisp Experiential Index (CCEI) (Crisp et al., 1978). Higher scores indicate more severe anxiety and scores of 9 or higher have previously been used to indicate more severe anxiety (Joinson et al., 2009). We dichtomised mothers into ‘anxious’ and ‘not anxious’ using this threshold. We used the mean of EPDS or CCEI scores at 18- and 32-weeks’ gestation to indicate antenatal depression or anxiety respectively.

### Negative controls: partner depression and anxiety

Mother’s partners completed the EPDS, validated for use in fathers (Edmondson et al., 2010) and CCEI at 18 weeks’ gestation. We used the same thresholds as above to derive binary variables for partner depression and anxiety during pregnancy.

### Outcomes: daytime wetting, bedwetting, soiling, and constipation at age 7 years

Mothers were asked ‘how often does your child: wet themselves during the day; wet the bed at night; and dirty their pants during the day’. Options included ‘never’; ‘occasional accidents but less than once a week’; ‘about once a week’; ‘2-5 times a week’; ‘nearly every day’, and ‘more than once a day’. We derived binary variables for daytime wetting, bedwetting, and soiling (occasional and more versus never). Mothers were also asked ‘has your child had any constipation in the past 12 months’: ‘yes and saw a doctor’; ‘yes but did not see a doctor’; ‘no did not have’. We derived a binary variable indicating any constipation versus none.

### Confounders

We adjusted for mother’s age at delivery, parity (‘1 or less’ or ‘2 or more’) and socioeconomic factors derived from responses to questionnaires during pregnancy including: occupational social class (using the 1991 British Office of Population and Census Statistics classification and dichotomised into ‘non-manual’ or ‘manual’); maternal education (‘A-level or degree’, ‘O level’, and ‘Certificate of Secondary Education/vocational qualification/none’); home ownership (‘owner or private renter’ and ‘renter or non-homeowner’); and a continuous score of financial difficulties. In models that included postnatal maternal psychopathology as the exposure, we additionally adjusted for child’s sex, gestation at birth (‘<32 weeks’, ‘32-36 weeks’, ‘37-41 weeks’, or ‘>=42 weeks’), and weight at delivery (‘< 2500g’, ‘2500-4499g’, or ‘>=4500g’).

### Statistical analysis

First, we used multivariable logistic regression models to examine associations between maternal psychopathology, separately in the antenatal and postnatal periods, and offspring incontinence/constipation (N=6,489). We examined models including both depression and anxiety to test for independent effects. Next, we examined whether exposure to antenatal or postnatal maternal psychopathology was more important in determining the risk of offspring incontinence/constipation by testing the fit of different life-course models to the data (see supplementary materials for further details). Lastly, we examined the evidence for a causal intra-uterine effect of antenatal maternal depression and anxiety on offspring incontinence/constipation using partner depression and anxiety assessed during the mother’s pregnancy as negative controls. The negative control analysis was conducted on a reduced sample (N=5,337) comprising participants with data available for both maternal and partner psychopathology assessed during pregnancy. We estimated the associations of maternal and partner depression and anxiety with offspring incontinence and constipation using multivariable logistic regression models adjusted for confounders. All analyses were conducted in Stata, version 17.

## Results

The analysis focused on 6,489 children with complete data on all maternal psychopathology exposures, childhood incontinence and constipation outcomes at age 7, and confounders. There were 512 (7.9%) children with daytime wetting, 1,009 (15.6%) with bedwetting, 446 (6.9%) with soiling, and 663 (10.2%) with constipation.

Table 1 shows the distribution of maternal and paternal psychopathology and confounders in the samples considered for this study. The restricted samples had lower proportions of antenatal maternal depression and anxiety, manual social class, home renters/non-owners, low maternal education, mothers with two or more pregnancies and children born prematurely or of low birthweight. There was little difference in the proportion of mothers with postnatal depression and anxiety across the samples. Proportions of partner depression and anxiety differed only slightly across the samples. There was little difference in mean maternal age at delivery or mean financial difficulties score across the samples and the sex distribution remained constant.

**Table 1.** Distribution of maternal and paternal psychopathology, child incontinence, and confounding factors in the samples considered for this study

|  | Data on maternal<br>antenatal EPDS & CCEI<br>N<=13,098 | Data on maternal<br>antenatal and postnatal<br>EPDS & CCEI<br>N<=9,272 | Data on maternal EPDS &<br>CCEI and continence<br>outcomes<br>N<=7,016 | Data on maternal EPDS &<br>CCEI, continence<br>outcomes, and<br>confounders<br>N=6,489 |
| --- | --- | --- | --- | --- |
| **Maternal Psychopathology** |  |  |  |  |
| Maternal Depression at 18/32<br>Weeks Gestation | 1,661 (12.7%) | 942 (10.2%) | 652 (9.3%) | 575 (8.9%) |
| Maternal Anxiety at 18/32 Weeks<br>Gestation | 1,952 (14.9%) | 1,197 (12.9%) | 837 (11.9%) | 742 (11.4%) |
| Maternal Depression at 33<br>Months | 1,131 (12.2%) | 1,128 (12.2%) | 820 (11.7%) | 745 (11.5%) |
| Maternal Anxiety at 33 Months | 1,397 (15.0%) | 1,393 (15.0%) | 1,003 (14.3%) | 911 (14.0%) |
| **Partner Psychopathology** |  |  |  |  |
| Partner Depression at 18 Weeks<br>Gestation | 392 (4.1%) | 248 (3.4%) | 188 (3.3%) | 176 (3.3%) |
| Partner Anxiety at 18 Weeks<br>Gestation | 481 (5.0%) | 335 (4.6%) | 246 (4.3%) | 232 (4.3%) |
| **Confounders** |  |  |  |  |
| Child's Sex (Male) | 6,728 (51.7%) | 4,766 (51.4%) | 3,588 (51.1%) | 3,318 (51.1%) |
| Manual Social Class (ref: Non-Manual) | 5,630 (49.0%) | 4,042 (45.9%) | 2,933 (43.3%) | 2,796 (43.1%) |
| Renter/Non-Homeowner (ref: Homeowner/Private Renter) | 2,327 (18.6%) | 1,280 (14.1%) | 804 (11.6%) | 693 (10.7%) |
| Parity – 2 or More (ref: 0 or 1) | 2,580 (20.3%) | 1,713 (18.8%) | 1,228 (17.8%) | 1,138 (17.5%) |
| Mean Financial Difficulties Score (SD) | 2.91 (3.54) | 2.68 (3.40) | 2.49 (3.29) | 2.47 (3.28) |
| Mean Maternal Age at Delivery (SD) | 28.12 (4.91) | 28.73 (4.66) | 29.11 (4.54) | 29.14 (4.48) |
| Maternal Education (reference category: A Level/Degree) |  |  |  |  |
| CSE/Vocational/None | 3,634 (30.0%) | 2,276 (25.1%) | 1,543 (22.3%) | 1,360 (21.0%) |
| O Level | 4,196 (34.7%) | 3,234 (35.6%) | 2,426 (35.1%) | 2,295 (35.4%) |
| Gestation at Birth (reference category: 37-41 Weeks) |  |  |  |  |
| <32 Weeks | 182 (1.4%) | 59 (0.6%) | 45 (0.6%) | 26 (0.4%) |
| 32-36 Weeks | 688 (5.3%) | 456 (4.9%) | 322 (4.6%) | 295 (4.6%) |
| ≥42 Weeks | 979 (7.5%) | 688 (7.4%) | 518 (7.4%) | 481 (7.4%) |
| Birth Weight (reference category: 2500-4499g) |  |  |  |  |
| <2500g | 720 (5.6%) | 425 (4.6%) | 300 (4.3%) | 262 (4.0%) |
| ≥4500g | 240 (1.9%) | 175 (1.9%) | 124 (1.8%) | 118 (1.8%) |

Abbreviations: SD, standard deviation. Maternal/paternal depression defined as an EPDS score of 13 or higher. Maternal/paternal anxiety defined as a CCEI score of 9 or higher. Child incontinence and constipation outcomes are defined as any versus no issue.

### Associations between maternal depression and anxiety in the antenatal and postnatal periods and offspring incontinence and constipation at age 7

The results in table 2 provide evidence that antenatal depression and anxiety were associated with increased odds of offspring constipation at age 7 after adjustment for confounders. The association between antenatal depression and constipation was attenuated after adjustment for antenatal anxiety, but the association with antenatal anxiety remained after adjustment for antenatal depression. There was no evidence of associations between antenatal depression or anxiety and daytime wetting, bedwetting or soiling in the adjusted models.

**Table 2.**
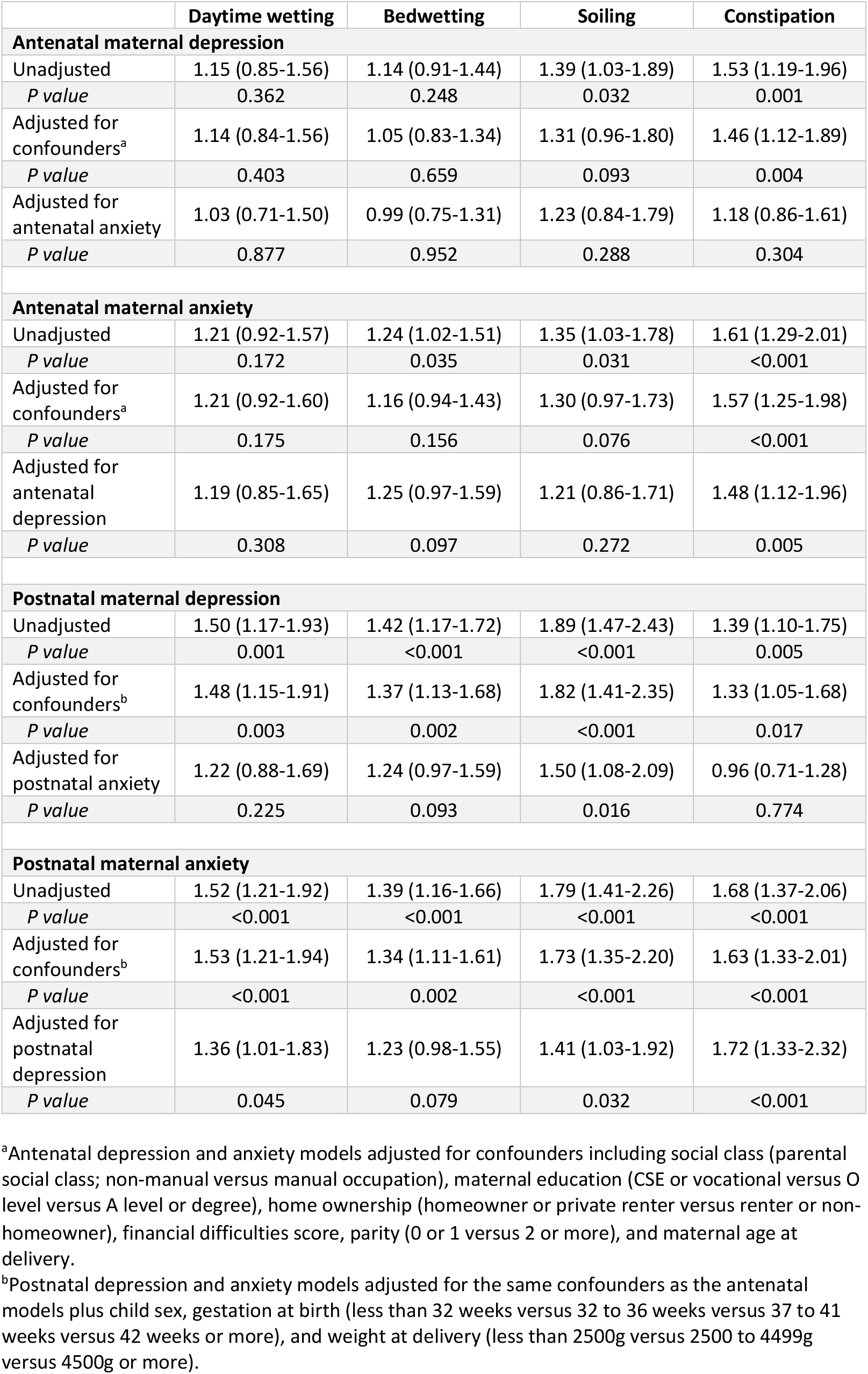
Odds ratios and 95% confidence intervals for the association between maternal depression and anxiety in the antenatal and postnatal periods and offspring daytime wetting, bedwetting, soiling, and constipation at age 7 years (N=6,489).

Postnatal depression and anxiety were associated with increased odds of daytime wetting, bedwetting, soiling, and constipation in the adjusted models. The associations with postnatal depression were attenuated following adjustment for postnatal anxiety, but there was still evidence for an association with soiling. The associations remained between postnatal anxiety and all outcomes, except bedwetting, after adjustment for postnatal depression.

### Life-course approach: testing for a critical/sensitive antenatal or postnatal period

Table 3 provides the adjusted odds ratios for the associations between the 4-level depression and anxiety exposure variables and the outcomes, and p values for the LR tests comparing the unconstrained and constrained models representing the different life-course hypotheses (supplementary table 1 presents the unadjusted analysis). There was evidence that the postnatal period was a critical period of exposure for maternal depression and anxiety and offspring daytime wetting, bedwetting, and soiling, but there was no support for an antenatal critical period model. Whilst there was some support for a sensitive antenatal period model in relation to these outcomes, the effect estimates for those exposed to maternal depression/anxiety only in the postnatal period were greater than the antenatal period, which is inconsistent with the antenatal period being sensitive. For constipation, there was some support for the antenatal period being a critical period of exposure for maternal depression; the critical postnatal period model was not supported, and the sensitive postnatal period model did not yield estimates consistent with that hypothesis. There was little support for a critical/sensitive antenatal or postnatal period model when examining the relationship between maternal anxiety and offspring constipation. Instead, there were increased odds of constipation for offspring exposed to maternal anxiety irrespective of the period of exposure.

**Table 3.**
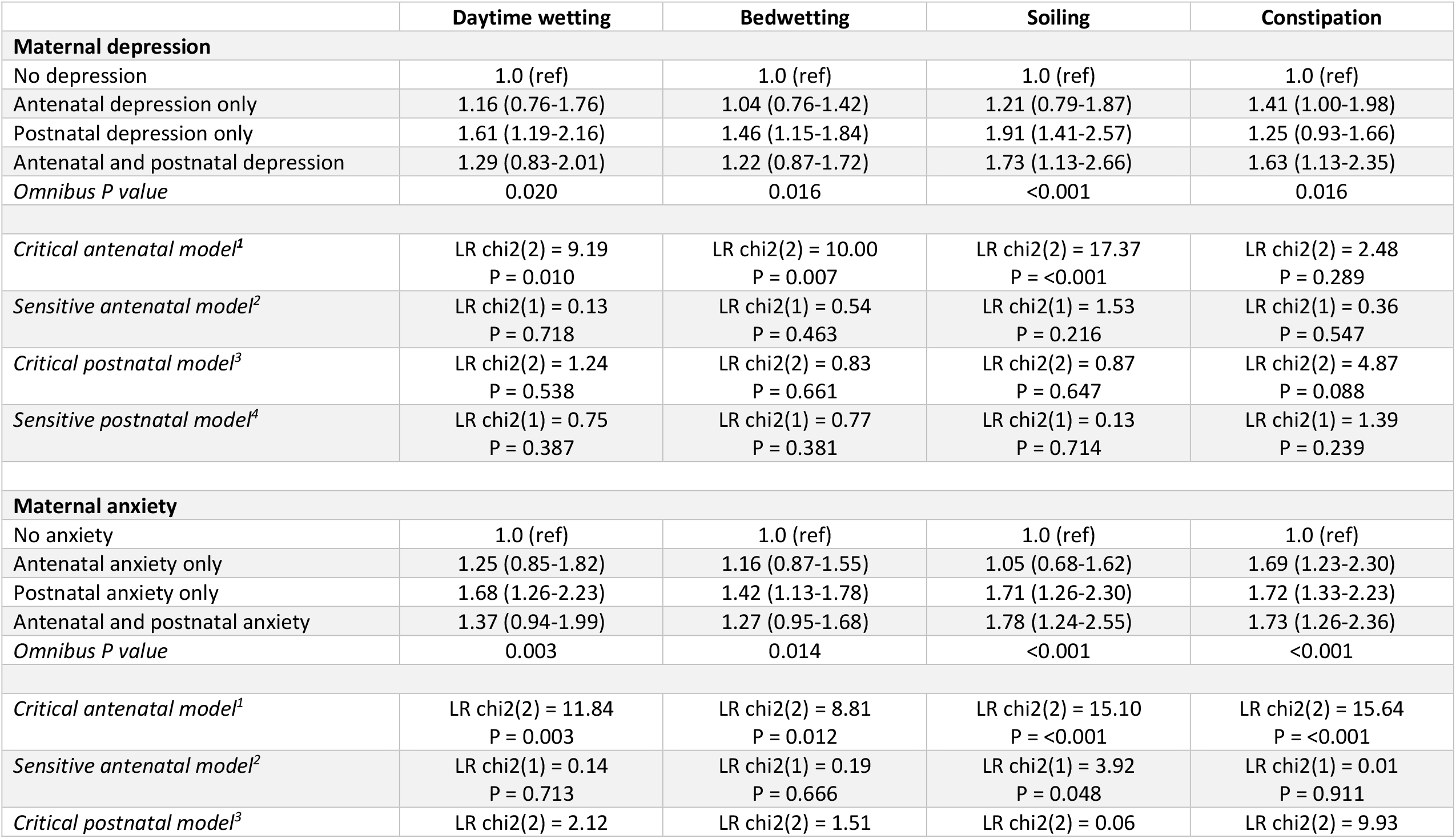
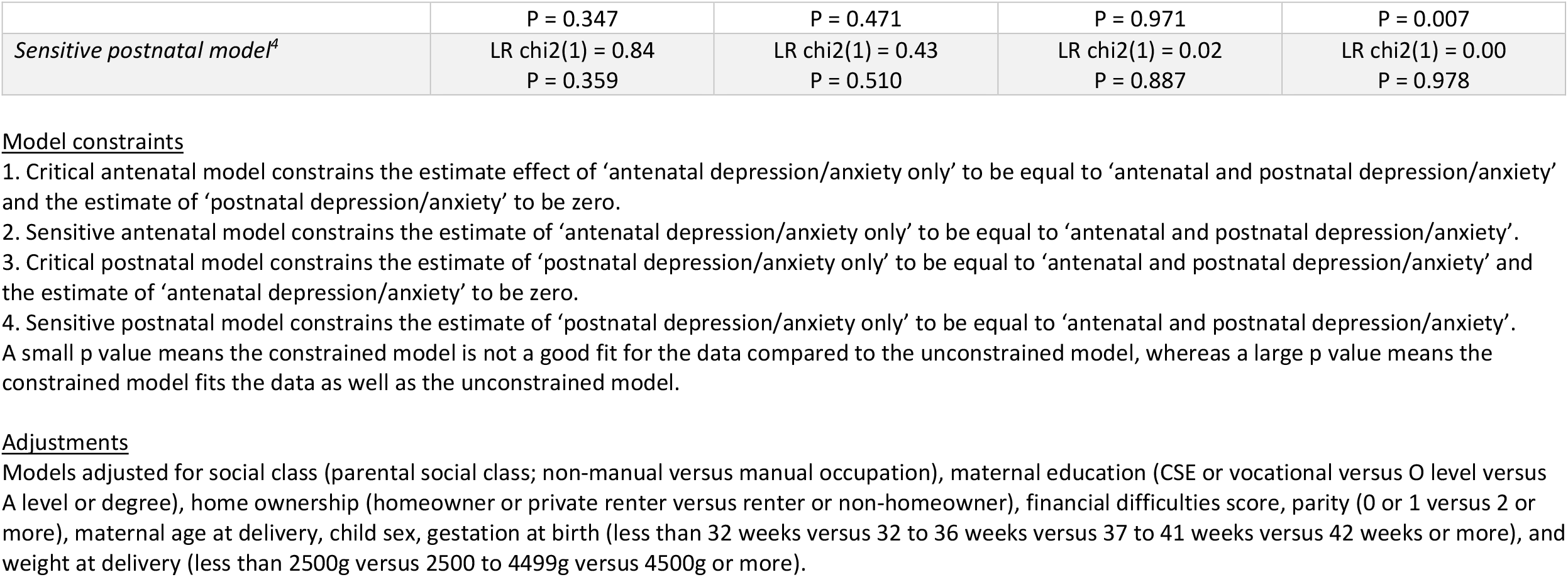
Adjusted odds ratios and 95% confidence intervals for the association between maternal depression and anxiety in the antenatal period only, postnatal period only, and both periods and offspring daytime wetting, bedwetting, soiling, and constipation (N=6,489)

### Negative control approach

Table 4 presents the adjusted odds ratios for the associations between maternal and partner depression and anxiety and offspring incontinence and constipation, with parents included in the same model (supplementary table 2 presents the unadjusted analysis and the analyses with parents included in separate models).

**Table 4.** Odds ratios and 95% confidence intervals for the association between maternal and paternal depression and anxiety in the antenatal period and offspring daytime wetting, bedwetting, soiling, and constipation, adjusted for confounders and with parents included in the same model (N=5,337).

|  | Daytime wetting |  | Bedwetting |  | Soiling |  | Constipation |  |
| --- | --- | --- | --- | --- | --- | --- | --- | --- |
|  | Maternal | Partner | Maternal | Partner | Maternal | Partner | Maternal | Partner |
| **Depression** |  |  |  |  |  |  |  |  |
| Adjusted | 1.20<br>(0.84-1.71) | 0.75<br>(0.40-1.41) | 0.90<br>(0.68-1.19) | 1.37<br>(0.93-2.00) | 1.26<br>(0.88-1.81) | 1.62<br>(1.00-2.64) | 1.54<br>(1.15-2.06) | 1.48<br>(0.96-2.27) |
| *P value* | 0.320 | 0.373 | 0.472 | 0.108 | 0.204 | 0.052 | 0.004 | 0.074 |
| **Anxiety** |  |  |  |  |  |  |  |  |
| Adjusted | 1.15<br>(0.84-1.58) | 1.09<br>(0.68-1.74) | 1.02<br>(0.80-1.29) | 1.32<br>(0.95-1.85) | 1.30<br>(0.95-1.78) | 1.72<br>(1.13-2.62) | 1.65<br>(1.28-2.12) | 1.49<br>(1.03-2.17) |
| *P value* | 0.389 | 0.734 | 0.881 | 0.099 | 0.107 | 0.011 | <0.001 | 0.036 |

Adjusted for social class (parental social class; non-manual versus manual occupation), maternal education (CSE or vocational versus O level versus A level or degree), home ownership (homeowner or private renter versus renter or non-homeowner), financial difficulties score, parity (0 or 1 versus 2 or more), and maternal age at delivery.

For constipation, there was a stronger association with maternal, compared with partner, depression. There was, however, considerable overlap in the 95% confidence intervals which is not consistent with a causal intrauterine effect. Both mother and partner anxiety were associated with offspring constipation. There was evidence for a stronger association between partner antenatal anxiety and offspring soiling. Maternal and partner depression and anxiety during pregnancy were not associated with daytime wetting or bedwetting.

## Discussion

We found evidence that exposure to maternal psychopathology in the postnatal period is associated with an increased risk of offspring incontinence and constipation at age 7. The associations with postnatal depression were attenuated following adjustment for postnatal anxiety, but the association between postnatal depression and soiling remained. There was evidence for an independent effect of maternal postnatal anxiety on offspring daytime wetting, soiling and constipation when models were adjusted for maternal postnatal depression. When we tested different life-course hypotheses, our results provided support for the postnatal, but not the antenatal, period being a critical period of exposure for maternal psychopathology in relation to offspring daytime wetting, bedwetting, and soiling. For constipation, there was some support for the antenatal period being a critical period of exposure for maternal depression. The risk of constipation in offspring exposed to maternal anxiety was increased irrespective of the period of exposure. There was little support for a causal intra-uterine effect of maternal antenatal psychopathology on offspring constipation. Instead, shared environmental or genetic factors provide a more likely explanation for the observed associations.

Major strengths of this study include the prospective design and the availability of data from a large birth cohort on validated measures of maternal depression and anxiety, and a wide range of confounders. Maternal depression and anxiety were assessed during pregnancy and postnatally, enabling us to examine whether these disorders have independent effects on offspring incontinence and constipation, and allowing us to test whether exposure to maternal psychopathology in the antenatal or postnatal period was more important. Paternal negative controls enabled us to account for measured and unmeasured confounding shared between mothers and partners, although it was not possible to account for any unmeasured confounding that was unique to mothers or partners.

Potential limitations of this study are the reliance on maternal reports of their child’s incontinence and constipation, and the lack of use of established diagnostic criteria. Whilst parents are likely to be aware of their child’s urinary and faecal incontinence, it is possible that some parents were unaware of their children’s constipation, especially if accompanied by soiling. The prevalence of constipation (10.2%) observed at age 7 in this cohort, however, is higher than the median prevalence (8.9%) reported in a systematic review of 0-to 18-year-olds (van den Berg et al., 2006), which suggests that constipation was not underreported. We chose to focus on any level of incontinence and constipation in this community sample, rather than on children who met currently established diagnostic criteria. There is evidence that children who experience incontinence and constipation below thresholds for clinical diagnosis still experience psychological distress (Joinson et al., 2006a). Furthermore, the demographic characteristics of ALSPAC participants limit generalisability of the findings to predominantly affluent, White UK populations. Further research is needed to determine whether the findings apply to more deprived populations and different ethnicities. The complete case sample used in this study was more socially advantaged than the original starting sample, but our estimates should be unbiased provided there are no systematic differences in the rates of the outcomes considered, after conditioning on the exposure and confounders included in the model (Hughes et al., 2019). Since our list of confounders is extensive and childhood incontinence in ALSPAC has been shown to be only weakly socially patterned (Butler and Heron, 2008), we believe this assumption is tenable in this current study.

Associations between maternal psychopathology and offspring incontinence and constipation have been reported in previous literature (Appak et al., 2017). However, such studies did not compare the effects of exposure to maternal psychopathology in the antenatal versus postnatal period, nor did they attempt to differentiate between the effects of maternal depression and anxiety. There is evidence that maternal anxiety and depression have different effects on offspring outcomes (Glover, 2014). This could be due to differential effects of distinct symptoms of anxiety (e.g. hyperarousal) and depression (e.g. low positive affect) on child development (Walker et al., 2020).

There are multiple plausible mechanisms that may underlie the associations between maternal psychopathology and incontinence observed in the current study. Maternal psychopathology is associated with increased hostility, more withdrawn, and less warm and positive parenting (National Research Council, 2009). Exposure to negative parenting behaviours around the time of potty training could disrupt the process of learning bladder and bowel control because mothers might be inconsistent or punitive in their approach and less responsive to the child’s toileting needs.

Maternal psychopathology is associated with increased stress in offspring (Ashman et al., 2022) and there is evidence that exposure to chronic stress can affect bladder and bowel functioning (Chess-Williams et al., 2021). Exposure to maternal psychopathology is also associated with an increased risk of offspring emotional and behavioural problems (Goodman et al., 2011), which are prospectively associated with incontinence and constipation (Joinson et al., 2019). There could also be a contribution of genetic factors, through shared genes predisposing mothers and offspring to psychopathology and/or incontinence and constipation. These environmental and genetic factors are also likely to explain the associations observed between antenatal maternal depression and constipation, because the negative control design found little evidence of a causal intra-uterine effect.

### Clinical implications

The current study highlights the importance of maternal mental health screening during the early years because children who were exposed to maternal postnatal psychopathology before their third birthday had a greater risk of incontinence and constipation at primary school age. Achieving bladder and bowel control are key developmental milestones, and failure to develop continence by primary school age can affect children’s mental health and quality of life. Our findings suggest that maternal postnatal anxiety has a stronger relationship with offspring incontinence/constipation.

Primary care health professionals should be vigilant to maternal psychopathology and its effects on child development, and parents should be offered support to deal with distress. There is also a need for evidence-based guidance to support parents to successfully navigate stressful developmental stages including potty training. Future research is required to examine whether the observed associations between maternal postnatal anxiety and offspring incontinence and constipation are causal and, if so, to examine underlying mechanisms that explain this association.

## Data Availability

All data in the present study are available subject to an approved application for data access from the ALSPAC Executive Committee.

## Abbreviations

ALSPAC: Avon Longitudinal Study of Parents and Children
CCEI: Crown Crisp Experiential Index
EPDS: Edinburgh Postnatal Depression Scale
HPA: Hypothalamic-Pituitary-Adrenal
LR: Likelihood Ratio

## Funding

The UK Medical Research Council and Wellcome Trust (Grant ref: 217065/Z/19/Z) and the University of Bristol provide core support for ALSPAC. This publication is the work of the authors and Gemma Sawyer, Jon Heron and Carol Joinson will serve as guarantors for the contents of this paper. This research was specifically funded by a Medical Research Council grant (ref: MR/V033581/1: Mental Health and Incontinence). Gemma Sawyer is supported by a Wellcome Trust PhD studentship in Molecular, Genetic and Lifecourse Epidemiology (ref: 218495/Z/19/Z). The authors have stated that they had no interests which might be perceived as posing a conflict or bias.

## Acknowledgements

We are extremely grateful to all the families who took part in this study, the midwives for their help in recruiting them, and the whole ALSPAC team, which includes interviewers, computer and laboratory technicians, clerical workers, research scientists, volunteers, managers, receptionists, and nurses.

## Notes

### Competing Interest Statement

The authors have declared no competing interest.

### Author Declarations

Ethics and Law Committee and the Local Research Ethics Committee of ALSPAC gave Ethical approval for this work (IRB00003312).

