## Supplementary Material for "The relationship between maternal psychopathology and offspring incontinence and constipation at school age: a prospective cohort study"

**Life-course approach: method of testing for a critical/sensitive antenatal or postnatal period**

We derived a 4-level maternal depression variable consisting of ‘no antenatal or postnatal depression’, ‘antenatal depression only’, ‘postnatal depression only’, and ‘depression in both periods’. We also derived a 4-level variable for maternal anxiety. We encoded an unconstrained model representing the relationship between antenatal and postnatal maternal depression/anxiety and the offspring outcomes as three parameter estimates (i.e. consistent with both time points and their interaction being associated with the outcome). We then encoded four lifecourse hypotheses as one or more constraints placed on this model: (a) *sensitive antenatal model*, which constrained the estimate of antenatal depression/anxiety to be equal to the estimate of depression/anxiety in both periods; (b) *sensitive postnatal model*, which constrained the estimate of postnatal depression/anxiety to be equal to the estimate of depression/anxiety in both periods; (c) *critical antenatal model*, constrained in the same way to (a) and, additionally, constrained the estimate of postnatal depression/anxiety to be zero; and (d) *critical postnatal model*, which constrained in the same way to (b) and, additionally, constrained the estimate of antenatal depression/anxiety to be zero. These models were adjusted for the confounders described previously. We used likelihood ratio (LR) tests to compare the constrained models to the unconstrained model. The unconstrained model will always provide the best fit to the data because it has most parameters. Therefore, a small p-value for the LR test provides evidence that the data are not consistent with the lifecourse hypothesis being examined (i.e. the fit of the constrained model is worse than the unconstrained model). A large p-value provides evidence that the considered lifecourse hypothesis could have given rise to the observed data.
